# Additional value of germline genomic scores beyond prostate-specific antigen alone for long-term and near-term prostate cancer risk

**DOI:** 10.64898/2026.11.17.26368323

**Authors:** Roshan Karunamuni, Joshua Stark, George Xu, Charles Brunette, Isla Garraway, Richard Hauger, Linda Kachuri, Adam Kibel, Michael A. Liss, Julie Lynch, Brenton Rose, Craig Teerlink, Jason L. Vassy, John Witte, VA Million Veteran Program, Tyler M. Seibert

**Author notes:** Corresponding author: Tyler Seibert. These authors contributed equally (Roshan Karunamuni, Joshua Stark, George Xu).

## Abstract

We evaluated whether germline genomic risk complements prostate-specific antigen (PSA) for prostate cancer early detection. P-CARE integrates a 601-variant polygenic hazard score, family history, and genetic ancestry. Long-term outcomes and clinically significant prostate cancer (csPCa) detection on biopsy were assessed in the Million Veteran Program (MVP), with external validation in the Prostate Cancer Prevention Trial (PCPT). Among 428,512 MVP participants, high versus low P-CARE risk was associated with increased hazards of treatment (HR 3.32) and metastasis (HR 3.32). Among men whose baseline PSA was measured at ages 40–49 with PSA 1–3 ng/mL, 15-year treatment/metastasis probabilities were 2.6%, 4.5%, and 10.2% across increasing P-CARE risk groups. High P-CARE risk also predicted csPCa on biopsy (OR 2.96), while PRS_PSA_ was inversely associated with outcomes. Findings were consistent in PCPT. Germline genomic risk complements measured PSA, supporting prospective evaluation of genetically informed screening strategies.

## Introduction

Prostate cancer (PCa) screening guidelines^1,2^ seek to balance the benefits of early detection against the harms of unnecessary testing, biopsy, overdiagnosis, and overtreatment. Decisions to initiate and continue screening are made through shared decision-making that considers age, co-morbidities, life expectancy, family history, ancestry, inherited cancer susceptibility, and patient preferences. Serum prostate-specific antigen (PSA) remains the principal initial screening test, with subsequent evaluation increasingly incorporating muti-parametric magnetic resonance imaging (MRI), and serum- or urine-based biomarkers to estimate the likelihood of clinically significant PCa and inform biopsy decisions^1–3^. Established risk factors (e.g., genetic ancestry, clinically actionable hereditary pathogenic genetic mutations, age, and family history of PCa) may further inform individualized screening decisions. Collectively, these approaches aim to improve detection of clinically significant or aggressive PCa while reducing unnecessary procedures.

Polygenic risk scores (PRS) have emerged as a promising approach to inform individualized PCa risk stratification. PRS estimates a man’s risk of developing PCa, by integrating contributions from numerous common genetic variants across the genome^4^. These risk scores have been used to quantify lifetime PCa risk^5–7^, predict lethal disease ^8^, and stratify risk across diverse and multi-ethnic cohorts^5,9–11^. More recently, Eeles *et al.* reported findings from the prospective BARCODE1 trial, in which men within the highest 10^th^ percentile of PRS risk underwent targeted PCa screening^12^. This genetically selected screening strategy identified clinically significant prostate cancers (csPCa) that would not have been detected using conventional PSA- or MRI-based screening approaches, suggesting that germline PRS may provide clinically relevant information beyond established screening measures.

Although polygenic risk is associated with PCa incidence and adverse clinical outcomes, an important unresolved question is whether genetic risk provides clinically relevant information after PSA is already known. Early baseline PSA is strongly associated with both near- and long-term PCa risk, raising the question of whether germline risk stratification meaningfully complements PSA after it has been measured^13^.

P-CARE (Prostate CAncer integrated Risk Evaluation) is a genomic risk model currently under study as an integral biomarker in the ProGRESS randomized trial (NCT05926102).^5^ In the present study, we evaluate whether P-CARE provides risk information complementary to measured PSA for both near-term detection of csPCa and longer-term PCa outcomes. We tested whether P-CARE was associated with detection of csPCa on biopsy in two cohorts: the Million Veteran Program (MVP; the dataset in which P-CARE was developed) and the Prostate Cancer Prevention Trial (PCPT; for independent validation). We additionally evaluated PRS_PSA_, a polygenic score for non-cancer-related PSA elevation, as a complementary genomic predictor that may further refine interpretation of measured PSA. Finally, given the long natural history of PCa, we examined associations between P-CARE and subsequent PCa outcomes to determine whether the genomic risk captured by P-CARE was also associated with clinically meaningful disease over longer-term follow-up. We hypothesized that P-CARE would provide risk information beyond PSA for estimating the probability of near- or long-term PCa risk.

## Methods

### Million Veteran Program (MVP)

Data from MVP were used to evaluate the utility of P-CARE in various screening scenarios. Clinical and phenotypic data—including age, PSA measurements, biopsy results, treatment dates, and dates of metastatic PCa diagnosis—were retrieved from the VA Corporate Data Warehouse using International Classification of Diseases (ICD) diagnosis codes and VA Central Cancer Registry data^14^. Details on MVP enrollment, data collection, genotyping, genetic imputation, and quality control have been previously described^15,16^.

P-CARE is a PCa model that integrates PHS601, a polygenic hazard score comprising of 601 common germline variants, with family history of PCa and genetic ancestry^5^. P-CARE estimates the relative age-dependent risk of developing PCa, including clinically significant, metastatic, and fatal disease. Participants were classified using pre-specified P-CARE thresholds as low risk (relative hazard <0.75), intermediate risk (relative hazard 0.75-1.5), or high risk (relative hazard >1.5).

We also adjusted for PRS_PSA_, a distinct polygenic score developed to estimate inherited predisposition to higher constitutive PSA levels in the absence of PCa^17^. A higher PRS_PSA_ indicates a genetic propensity toward PSA elevation in the absence of PCa. PRS_PSA_ was analyzed as a continuous variable standardized to a mean of 0 and standard deviation of 1. For both polygenic models, previously published variants, weights, and locked model definitions were applied. Variant dosages (the estimated number of copies of each genetic variant ranging from 0 to 2) were extracted from genotype data imputed using the TOPMed reference panel. To display joint stratification by P-CARE and PRS_PSA_ in Figure 2B, biopsy records were categorized into combined genetic risk groups using the definitions in Supplementary Table 11.

#### MVP Long-term Risk Analysis

Because PCa treatment reduces risk of metastasis^18^, the *long-term risk* analysis evaluated the association between information available at a participant’s first recorded PSA measurement (baseline PSA) and subsequent transition to PCa treatment or metastatic disease, in a multi-state model. Men were eligible if their first PSA on record was collected before any PCa diagnosis or treatment; between the ages of 40 and 70 years; and between a value of 0.1 and 20 ng/ml.

Transition to PCa treatment was defined as receipt of androgen deprivation therapy (ADT), radiotherapy, or prostatectomy. Diagnosis of metastatic PCa was considered separately as an additional clinical endpoint. Men who did not transition to treatment or develop metastatic PCa were censored at time of last follow-up.

We then constructed a multi-state model with three states (screening, treatment, and metastasis) and two transitions: screening-to-treatment and screening-to-metastasis. Within the model, time was recorded as years since baseline PSA measurement. Model covariates included previously defined P-CARE risk groups (low, intermediate, high)^5^, baseline PSA value (ng/ml), age at baseline PSA (years), genetic ancestry (European, African, Other), and PRS_PSA_ (standardized to mean of 0 and standard deviation of 1). Patients were censored at time of last follow-up or death. A secondary analysis was restricted to men whose baseline PSA was measured at ages 40–49 to examine long-term risk stratification after an early midlife PSA measurement. Ninety-five percent confidence intervals for all HRs were calculated using the Wald method. In this subgroup, we used the fitted multistate model to estimate 15-year transition probabilities across combinations of baseline PSA and P-CARE risk group.

#### MVP Near-term Risk Analysis

The second analysis investigated the *near-term risk* of finding PCa on biopsy. Among men with a PCa diagnosis, we identified prostate biopsies (CPT codes: 55700, 55705, or 55706) performed on or before their initial date of PCa diagnosis. Among those without a recorded PCa diagnosis, all biopsies in the follow-up period were included. Biopsies were eligible if a PSA measurement between 0.1 and 20 ng/ml was available within the preceding 12 months. For those with a PCa diagnosis, the outcome of the biopsy was determined by cross-referencing the dataset with Gleason scores derived from natural language processing of pathology reports^5^.

Each biopsy was assigned the Gleason score of the temporally closest pathology report occurring within 6 months after the date of biopsy. csPCa was defined as grade group (GG) ≥2. Benign biopsies were detected by identifying biopsy events in the medical record without a previous or subsequent PCa diagnosis.

We then fit a logistic regression model using the biopsy result (csPCa vs. benign or GG1) as the outcome variable. Model predictor variables included: P-CARE risk group (low, intermediate, high), age at biopsy (years), genetic identified ancestry (European, African, Other), PRS_PSA_ (standardized), and an indicator to identify those instances where a biopsy had already been performed (i.e., benign or GG1). Cluster-robust standard errors were estimated to account for within-cluster correlation of the small subset of patients who provided more than one biopsy to the dataset.

### Prostate Cancer Prevention Trial (PCPT)

Data from Prostate Cancer Prevention Trial (PCPT) were used as an independent validation cohort to assess the generalizability of the findings from the MVP *long-term* and *near-term risk* analyses. The PCPT was a large, randomized clinical trial that enrolled 18,882 men aged ≥55 years to evaluate finasteride versus placebo for PCa prevention.^19^ It included standardized longitudinal PSA screening and protocol-driven biopsy, providing a well-characterized cohort for external validation. Importantly, PCPT participants were not included in the genome-wide association (GWAS) studies used to derive either P-CARE or PRS_PSA_. Phenotypic data (age at baseline PSA, treatment arms, baseline PSA, PCa diagnosis, and negative biopsy results) were obtained from the trial consortium. Genotype data were available for 6,054 PCPT participants, who were genotyped on the Illumina Infinium Global Screening Array 24 v2.0. These data were imputed using the 1000 Genomes Phase 3 reference panel, and variant dosages were extracted for analysis.

#### PCPT Long-term Risk Analysis

For the PCPT *long-term risk* analysis, the primary outcome was defined as the number of days between enrollment and any PCa diagnosis (chosen because it was the trial’s primary outcome). Participants without a PCa diagnosis were censored at last follow-up or death. A multivariable Cox proportional hazards model was fit using the time to PCa diagnosis as the outcome, and the following predictor variables: P-CARE risk group (low, intermediate, high), age at baseline (years), baseline PSA (ng/ml), treatment group (placebo, finasteride), PRS_PSA_ (scaled) and genetic ancestry.

A secondary analysis evaluated time to diagnosis of csPCa, defined as before. Participants without a PCa diagnosis were censored at last follow-up or death, whereas participants with GG1 cases were censored at date of diagnosis, as most received radical treatment in that era.

#### PCPT Near-term Risk Analysis

The *near-term risk* outcome was defined as the probability of a positive result on biopsy within 12 months of PSA result. We then fit a logistic regression model using the following predictor variables: P-CARE risk group (low, intermediate, high), age at biopsy (years), pre-biopsy PSA (ng/ml), treatment group (placebo, Finasteride), PRS_PSA_ (scaled), and genetic ancestry.

### Software tools

Extraction, manipulation, and filtering of genetic data were performed in PLINK (v.2.0.0). Estimation of multi-state models, Cox proportional hazards models, and logistic regression models were performed in R (v. 4.4.2). Values were obtained using the “msfit” and “probtrans” functions in the “mstate” package in R.

## Results

Characteristics of the cohorts used in the *long-term* and *near-term risk* analyses for MVP and PCPT are presented in Table 1. In total, 428,512 men met criteria for inclusion in the *long-term risk* analyses in MVP, with 57,706 biopsies from 46,652 men available for *near-term risk* analysis. From PCPT, longitudinal data from 6,054 men were available for long-term analysis and 4,308 biopsies from 4,269 men were included in near-term analyses. Among men who remained untreated and metastasis-free, median age at last follow-up was 72.6 years (IQR 65.8–77.2) in MVP and 83.0 years (IQR 79.7–86.9) in PCPT.

**Table 1.** Characteristics of the Million Veteran Program (MVP) and Prostate Cancer Prevention Trial (PCPT) analytic cohorts for long-term PCa risk and near-term biopsy risk analyses. Values are n (%) or median [interquartile range], as applicable.

| **Characteristic** | **Long-term risk analysis in MVP** | **Long-term risk analysis in PCPT** | **Near-term risk analysis in MVP** | **Near-term risk analysis in PCPT** |
|---|---|---|---|---|
| Number of men | 428,512 | 6,054 | 46,652 | 4,269 |
| treatment | 44,092 (10.3%) |  |  |  |
| metastatic PCa | 6,774 (1.58%) |  |  |  |
| P-CARE risk groups |  |  |  |  |
| low | 96,068 (22.4%) | 1,810 (29.9%) | 5,546 (11.9%) | 1,303 (30.2%) |
| intermediate | 157,522 (36.8%) | 2,760 (45.6%) | 13,906 (29.8%) | 1,955 (45.4%) |
| high | 174,922 (40.8%) | 1,484 (24.5%) | 27,200 (58.3%) | 1,050 (24.4%) |
| Genetic Ancestry |  |  |  |  |
| European | 300,613 (70.2%) |  | 29,993 (64.3%) |  |
| African | 84,089 (19.6%) |  | 12,440 (26.7%) |  |
| Other | 43,810 (10.2%) |  | 4,219 (9.04%) |  |
| Median age [years] at: |  |  |  |  |
| baseline PSA | 56.4 [50.7-62.1] | 63 [59-67] |  |  |
| treatment | 67.8 [62.6-72.8] |  |  |  |
| metastatic PCa | 72.4 [67.1-77.5] |  |  |  |
| PCa diagnosis |  | 71 [66.5-75.2] |  |  |
| last follow-up | 72.6 [65.8-77.2] | 83 [79.7-86.9] |  |  |
| baseline PSA [ng/ml] | 0.9 [0.5-1.5] | 1.1 [0.6-1.6] |  |  |
| Number of biopsies |  |  | 57,706 | 4,308 |
| positive result |  |  | 14,896 (25.8%) | 70 (1.62%) |
| Age at biopsy [years] |  |  | 65.2 [60.5-69.6] | 68.9 [65.0-73.1] |
| PSA pre-biopsy |  |  | 5.7 [4.4-7.8] | 0.9 [0.4-1.8] |
| PCa diagnosis |  | 368 (6.08%) |  |  |
| treatment group |  |  |  |  |
| placebo |  | 2,977 (49.2%) |  | 2,115 (49.1%) |
| Finasteride |  | 3,077 (50.8%) |  | 2,193 (50.9%) |

### MVP Long-term Risk

HRs for screening-to-treatment and screening-to-metastasis transitions in the *long-term risk* analysis are presented in Table 2 for the full MVP cohort and for the subgroup of men whose baseline PSA was obtained at age 40-49 years, as early baseline PSA has known prognostic value and is endorsed by guidelines^2,13^. To illustrate risk across combinations of baseline PSA and P-CARE risk, we estimated the 15-year probabilities of transitioning from screening-to-treatment or screening-to-metastasis among men whose baseline PSA was obtained in their 40s (Figure 1). Predicted probabilities are shown for combinations of P-CARE risk group and baseline PSA value, with 95% confidence intervals represented by the vertical extent of each color-coded rectangle.

**Figure 1.**
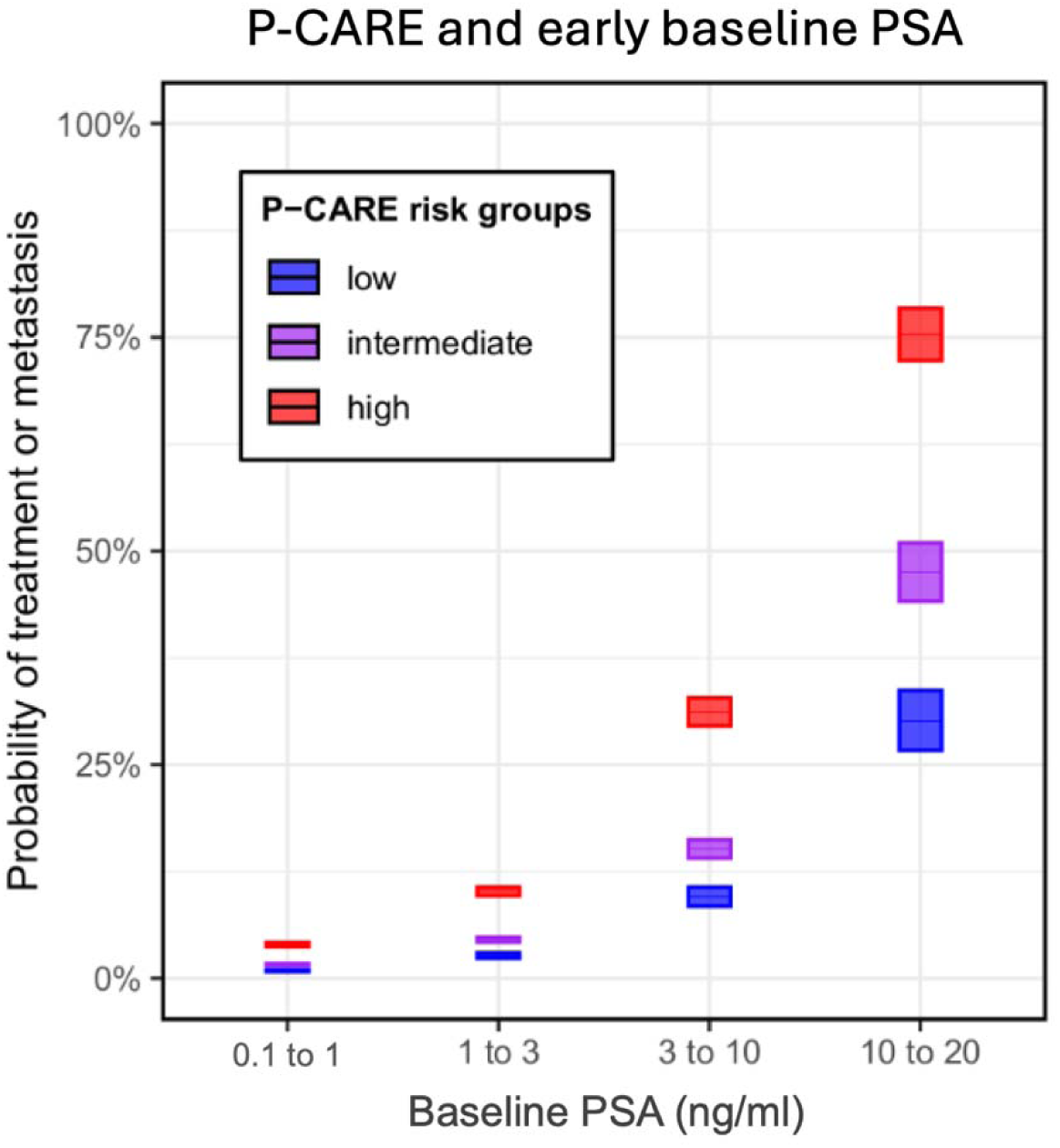
Probability of transitioning to treatment or metastasis 15 years after baseline as a function of the baseline PSA value. Visualization is limited to men who were first screened in their forties. Color-coded rectangles represent the 95% confidence interval for combinations of P-CARE risk group and PSA value. Transition probabilities were estimated from the multistate model using the ‘probtrans’ function in the ‘mstate’ R library. Additional data in Supplementary Table 12.

**Table 2.**
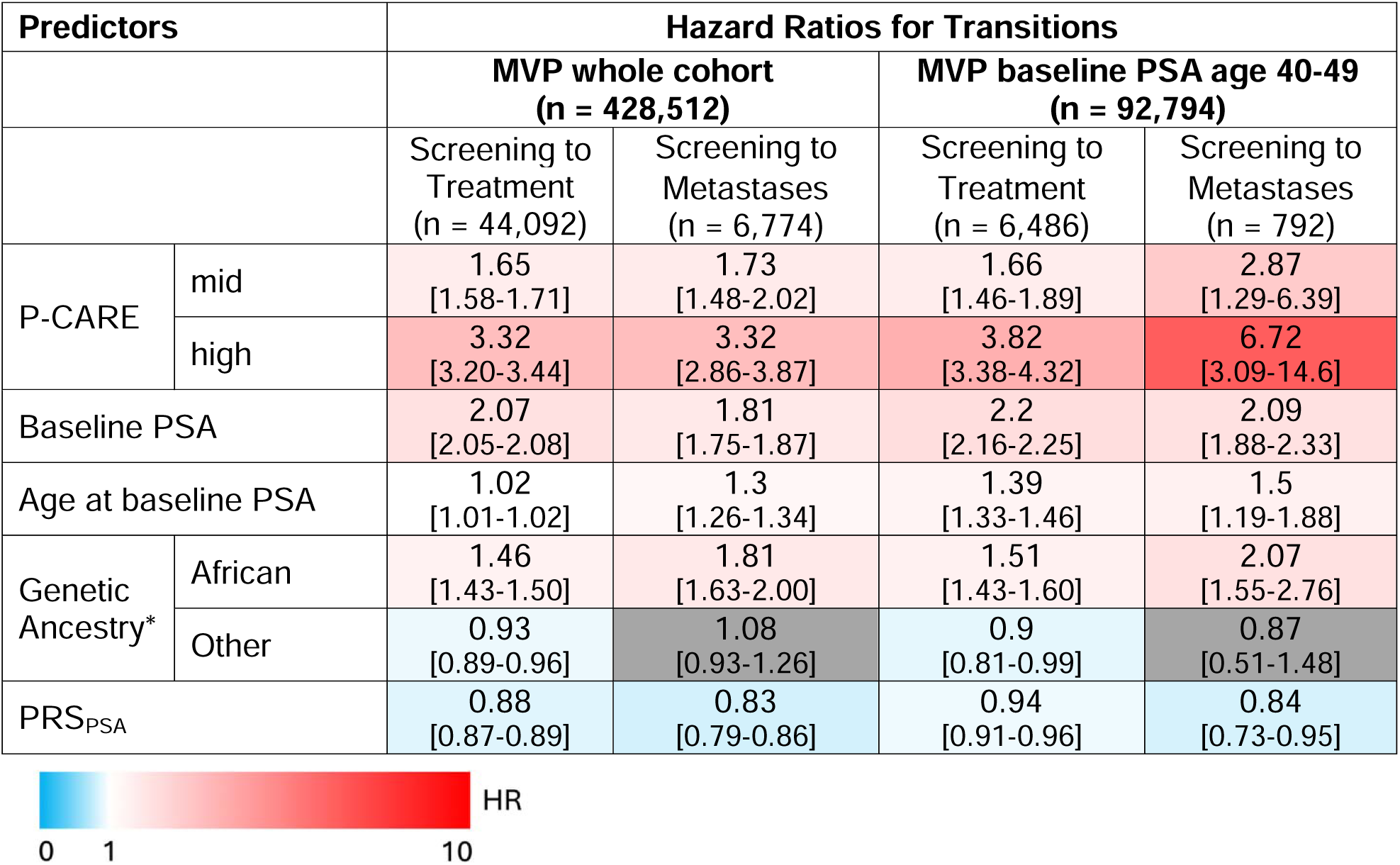
Hazard ratios for screening-to-treatment and screening-to-metastasis transitions in the long-term risk analysis of the full MVP cohort and the subgroup of men whose baseline PSA was obtained in their forties. 95% confidence intervals are tabulated in brackets. Values for baseline PSA are for every doubling in value; age at baseline PSA for every increase in 5 years; PRS_PSA_ for every increase in 1 standard deviation. Gray boxes denote no statistical significance. * European ancestry is reference

| **Predictors** |  | **Hazard Ratios for Transitions** |  |  |  |
|---|---|---|---|---|---|
|  |  | **MVP whole cohort<br>(n = 428,512)** |  | **MVP baseline PSA age 40-49<br>(n = 92,794)** |  |
|  |  | Screening to Treatment<br>(n = 44,092) | Screening to Metastases<br>(n = 6,774) | Screening to Treatment<br>(n = 6,486) | Screening to Metastases<br>(n = 792) |
| P-CARE | mid | 1.65<br>[1.58-1.71] | 1.73<br>[1.48-2.02] | 1.66<br>[1.46-1.89] | 2.87<br>[1.29-6.39] |
|  | high | 3.32<br>[3.20-3.44] | 3.32<br>[2.86-3.87] | 3.82<br>[3.38-4.32] | 6.72<br>[3.09-14.6] |
| Baseline PSA |  | 2.07<br>[2.05-2.08] | 1.81<br>[1.75-1.87] | 2.2<br>[2.16-2.25] | 2.09<br>[1.88-2.33] |
| Age at baseline PSA |  | 1.02<br>[1.01-1.02] | 1.3<br>[1.26-1.34] | 1.39<br>[1.33-1.46] | 1.5<br>[1.19-1.88] |
| Genetic Ancestry* | African | 1.46<br>[1.43-1.50] | 1.81<br>[1.63-2.00] | 1.51<br>[1.43-1.60] | 2.07<br>[1.55-2.76] |
|  | Other | 0.93<br>[0.89-0.96] | 1.08<br>[0.93-1.26] | 0.9<br>[0.81-0.99] | 0.87<br>[0.51-1.48] |
| PRS<sub>PSA</sub> |  | 0.88<br>[0.87-0.89] | 0.83<br>[0.79-0.86] | 0.94<br>[0.91-0.96] | 0.84<br>[0.73-0.95] |

In the full cohort, men in the intermediate- and high-risk P-CARE groups had higher hazards of subsequent treatment (HR 1.65, 95% CI 1.58-1.71 and HR 3.32, 95% CI 3.20-3.44, respectively) and metastatic disease (HR 1.73, 95% CI 1.48-2.02 and HR 3.32, 95% CI 2.86-3.87, respectively). Higher baseline PSA was also associated with both outcomes, whereas higher PRS_PSA_ was inversely associated with treatment and metastatic disease (HR 0.88, 95% CI 0.87-0.89 and HR 0.83, 95% CI 0.79-0.86, respectively).

Among men whose baseline PSA was obtained at ages 40-49, P-CARE remained associated with both outcomes. Compared with the low-risk group, men in the intermediate and high P-CARE group had a higher hazard of treatment (HR 1.66, 95% CI 1.46-1.89 and HR 3.82, 95% CI 3.38–4.32, respectively) and metastatic disease (HR 2.87, 95% CI 1.29-6.39 and HR 6.72, 95% CI 3.09–14.6). In this early midlife group, 15-year probabilities of transition to treatment or metastasis for those with baseline PSA 1–3 ng/mL were 2.6%, 4.5%, and 10.2% for the low-, intermediate-, and high-risk groups, respectively; for baseline PSA 3–10 ng/mL, the corresponding probabilities were 9.6%, 15.1%, and 31.2% (Figure 1, Supplementary Table 12).

### MVP Near-term Risk of Positive Biopsy

Odds ratios (ORs) for the *near-term risk analysis* are presented in Table 3. Similar results were found when restricting analyses to the first available biopsy for each man (Supplementary Table 5). To illustrate risk across combinations of P-CARE risk group and pre-biopsy PSA, we estimated the average predicted probabilities of detecting csPCa on biopsy (Figure 2A). Figure 2B similarly displays predicted probabilities using genetic risk groups defined by combining P-CARE risk group with PRS_PSA_ percentile (see Supplementary Table 11 for additional details).

**Table 3.** Odds ratio for detection of csPCa on biopsy in the MVP risk analysis. 95% confidence intervals are tabulated in square brackets. Values for PSA pre-biopsy are for every doubling in value; age at biopsy for every increase in 5 years; PRS_PSA_ for every increase in 1 standard deviation. Gray boxes denote no statistical significance. * European ancestry is reference.

| **Predictors** |  | **OR for positive biopsy result (csPCa, n = 14,896)** | **OR for positive biopsy result (GG≥3, n = 7,522)** |
|---|---|---|---|
| P-CARE | mid | 1.62<br>[1.49-1.76] | 1.44<br>[1.29–1.60] |
|  | high | 2.96<br>[2.74-3.21] | 2.18<br>[1.97-2.41] |
| PSA pre-biopsy |  | 1.97<br>[1.92-2.03] | 2.2<br>[2.12-2.29] |
| Age at biopsy |  | 1.27<br>[1.25-1.29] | 1.34<br>[1.31-1.37] |
| Genetic Ancestry | African | 1.48<br>[1.41-1.55] | 1.42<br>[1.33-1.50] |
|  | Other | 0.94<br>[0.87-1.02] | 0.99<br>[0.89-1.09] |
| Previous biopsy |  | 0.23<br>[0.21-0.24] | 0.25<br>[0.23-0.27] |
| PRS<sub>PSA</sub> |  | 0.72<br>[0.70-0.73] | 0.75<br>[0.73-0.77] |

OR
0 1 10

**Figure 2.**
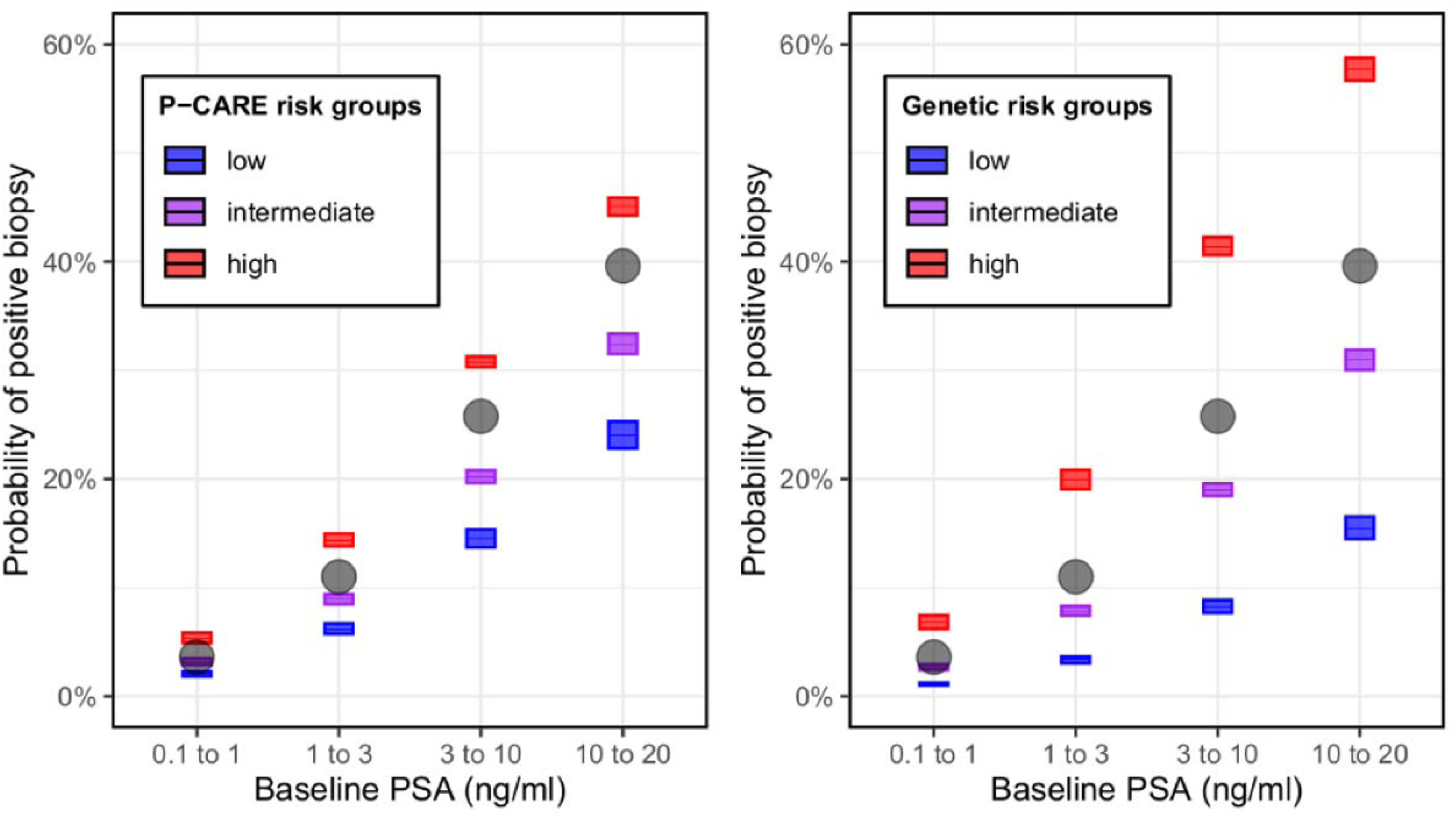
(A) Predicted probability of a positive biopsy for various combinations of P-CARE risk group and pre-biopsy PSA values. Gray circles represent the averaged values at that PSA level. Low (n = 7,006), Intermediate (n = 17,468), High (n = 33,232). (B) Probabilities for combinations of genetic risk group, combining both P-CARE and PRS_PSA_, and pre-biopsy PSA. Low (n = 896), Intermediate (n=8,931), High (n=6,315). Color-coded rectangles represent the predicted probabilities for each combination of P-CARE risk group and pre-biopsy PSA, with the vertical extent representing the corresponding 95% confidence interval. Gray circles represent the average predicted probability based on pre-biopsy PSA value alone.

Men in the intermediate- and high-risk P-CARE groups had higher odds of csPCa on biopsy (OR 1.62, 95% CI 1.49–1.76; and OR 2.96, 95% CI 2.74–3.21, respectively). Each doubling of pre-biopsy PSA was also associated with higher odds of csPCa (OR 1.97, 95% CI 1.92–2.03), whereas higher PRS_PSA_ was inversely associated with csPCa (OR 0.72 per standard deviation, 95% CI 0.70–0.73). These associations were similar when GG≥3 disease was used as the biopsy outcome (Table 3).

### PCPT Long-term Risk and Near-term Risk

HRs for the *long-term risk* analysis and ORs for the *near-term risk* analysis in PCPT are presented in Table 4. Results of the secondary analysis using the time to diagnosis of csPCa are tabulated in Supplementary Table 6.

**Table 4.** Hazard ratios for f time from baseline to diagnosis of PCa in PCPT, as well as odds ratio for near-term risk analysis in PCPT (csPCa). 95% confidence intervals are tabulated in square brackets. Values baseline PSA are for every doubling in value; age baseline for every increase in 5 years; PRS_PSA_ for every increase in 1 standard deviation. Gray boxes denote no statistical significance.

| **Predictors** |  | ***Long-term risk*** **HR for PCa diagnosis (n = 71)** | ***Near-term risk*** **OR for positive biopsy result (csPCa, n = 70)** | ***Near-term risk*** **OR for positive biopsy result (GG≥3, n = 30)** |
|---|---|---|---|---|
| P-CARE | mid | 1.61<br>[1.20-2.16] | 1.87<br>[0.92-4.11] | 1.78<br>[0.60-6.53] |
|  | high | 3.03<br>[2.24-4.11] | 4.41<br>[2.17-9.73] | 4.13<br>[1.42-15.0] |
| Baseline PSA |  | 1.52<br>[1.34-1.72] | 2.22<br>[1.78-2.77] | 2.41<br>[1.75-3.39] |
| Age at baseline |  | 1.1<br>[1.00-1.20] | 1.43<br>[1.16-1.75] | 1.34<br>[0.98-1.82] |
| PRS<sub>PSA</sub> |  | 0.81<br>[0.73-0.91] | 0.69<br>[0.53-0.89] | 0.83<br>[0.55-1.24] |
| Treatment group (Finasteride) |  | 0.59<br>[0.48-0.73] | 1.65<br>[0.98-2.76] | 2.65<br>[1.23-5.78] |
| Race (other) |  | 1.19<br>[0.75-1.90] | 2.07<br>[0.69-4.97] | 4<br>[1.13-10.9] |

OR or HR
0 1 10

Findings were consistent with those observed in MVP. Men in the intermediate- and high-risk P-CARE groups had higher hazards of subsequent PCa diagnosis (HR 1.61, 95% CI 1.20–2.16; and HR 3.03, 95% CI 2.24–4.11, respectively). Higher baseline PSA was also associated with PCa diagnosis (HR 1.52 per doubling, 95% CI 1.34–1.72), whereas higher PRS_PSA_ was inversely associated with diagnosis (HR 0.81 per standard deviation, 95% CI 0.73–0.91).

In the near-term analysis (PCa diagnosed on biopsy within 12 months of PSA result), the intermediate-risk group had an OR of 1.87 (95% CI 0.92–4.11), and the high-risk group had an OR of 4.41 (95% CI 2.17–9.73). Each doubling of pre-biopsy PSA was associated with higher odds of csPCa disease (OR 2.22, 95% CI 1.78–2.77), whereas higher PRS_PSA_ was inversely associated with csPCa detection (OR 0.69 per standard deviation, 95% CI 0.53–0.89).

## Discussion

In this study, we found that germline polygenic risk is independently associated with both long-term PCa outcomes and near-term detection of clinically significant disease after accounting for PSA and other clinical covariates. Prior studies have demonstrated that polygenic risk scores (PRS) can stratify lifetime PCa risk and identify men who may derive more or less benefit from PCa screening^5,11,14,20,21^. Our findings extend this literature by suggesting genomic risk stratification may also have a role in guiding PCa screening and biopsy decisions in men who have already had a PSA test, suggesting polygenic scores might have utility at multiple points along the early-detection pathway.

The *MVP long-term risk* analyses demonstrated that men in the intermediate- and high-risk P-CARE groups were, on average, 1.6 to 3 times more likely to require treatment or be diagnosed with metastatic PCa at any time after the initial PSA test, compared to men in the low germline genomic risk group. The *MVP near-term risk* analyses demonstrated that men undergoing biopsy who were in the intermediate and high P-CARE groups were 1.7 to 3 times more likely to have a csPCa on biopsy than those in the low germline genomic risk group. Effect sizes were consistent across the two datasets, suggesting a robust association between P-CARE and outcomes within the setting of PCa screening.

One guideline-concordant strategy to risk-stratify future screening decisions is to obtain an early baseline PSA in early midlife^2,22^. The *long-term* risk analysis suggests P-CARE is complementary to early baseline PSA, as P-CARE was independently associated with PCa outcomes even in those men with first PSA in their forties. As illustrated in Figure 1, P-CARE stratifies long-term risk for men in their forties across various ranges of early baseline PSA values. While the overall risk for a man in his forties with a baseline PSA <1 ng/ml is low, the data suggest that men in the high P-CARE group might have a similar long-term risk to those with low or intermediate P-CARE who have baseline PSA values between 1 and 3 ng/ml. A similar conclusion can be reached for PSA >3 ng/ml. This suggests that men in the high P-CARE risk group may benefit from earlier and more frequent screening to align their pathways with men who present with higher baseline PSA values.

Likewise, the *near-term risk* analysis suggests P-CARE groups can be informative for biopsy decisions. Figure 2 shows that men in the high P-CARE risk group with lower PSA levels have approximately the same rate of csPCa on biopsy as those in the low or intermediate P-CARE groups with higher PSA values. The effect is more pronounced when considering the combined risk stratification of P-CARE and PRS_PSA_, especially for men with high PSA. In this case, men in the high combined genetic risk groups had approximately the same rate of positive biopsies as all men with higher PSA values. A consolidated genetic model combining the effects of P-CARE and PRS_PSA_ may show substantial benefit in identifying those men most at risk and thus most likely to gain from a biopsy, as well as reducing overdiagnosis in men with low genetic risk who may present with PSA elevation. Furthermore, results from the BARCODE1 trial^12^ showed that men in the upper 10 percent of PRS distribution had a higher rate of clinically significant findings on biopsy compared to using PSA alone. While the definitions of high genetic risk groups in the two analyses are different, our findings corroborate that men with high inherited risk have a higher risk of csPCa on biopsy than is appreciated when only using PSA. On the other end of the risk spectrum, men in the low P-CARE risk group with intermediate levels of PSA (1.0 to 10 ng/ml) have comparable rates of positive biopsy to those with PSA levels less than 1.0 ng/ml.

This suggests that combining genetic risk scores with other clinical information could help identify men who might safely avoid biopsy or who should undergo additional biomarker tests before a biopsy is performed.

These findings argue against the hypothesis that baseline PSA fully captures clinically relevant PCa risk. Rather than serving as interchangeable risk markers, baseline PSA and germline markers (P-CARE and PRS_PSA_) appear to provide complementary information across both near-and long-term time points.

Because P-CARE was developed to estimate overall PCa susceptibility rather than tumor aggressiveness specifically, associations with clinically significant or metastatic PCa should be interpreted in this context. Men with greater underlying susceptibility to PCa will, by definition, also have greater absolute risk of developing clinically significant disease. Thus, these associations do not necessarily indicate that P-CARE preferentially identifies biologically aggressive cancers independent of its association with overall PCa risk. Importantly, a risk-stratified screening strategy does not necessarily require preferential prediction of aggressive tumor biology to have clinical utility. Enriching screening among men with greater underlying PCa susceptibility may also enrich for the absolute number of clinically significant cancers detected. However, such an approach may concurrently increase the detection of low-grade disease, emphasizing the importance of minimizing downstream overtreatment through appropriate use of contemporary diagnostic evaluation (e.g., MRI), risk stratification, and active surveillance^2,23–25^.

The findings of this study must be taken in context of its limitations. MVP is an exceptionally rich dataset in terms of both size and diversity. However, the clinical outcomes were extracted retrospectively from a combination of registries and medical reports, which may introduce a degree of uncertainty into the ages at which men experienced these outcomes. Because the near-term analysis relied on biopsy procedures and Gleason scores captured in VA records, biopsies performed outside the VA—particularly among Medicare-eligible veterans—may have been missed; when subsequent treatment was recorded at the VA, the cancer diagnosis could be captured without the corresponding biopsy-level Gleason outcome, potentially leading to underascertainment of biopsy-detected csPCa. P-CARE was originally developed using data from MVP, although for a different endpoint, and MVP therefore does not constitute a fully independent validation cohort. Because the published P-CARE score and risk categories were applied without refitting, the present analysis is less susceptible to model overfitting; nevertheless, reuse of the development population may result in optimistic estimates of its association with the outcomes evaluated here. We addressed these concerns by applying P-CARE in PCPT, an independent cohort derived from a prospective randomized clinical trial, where we observed comparable associations. That noted, PCPT is a highly selected population with low PSA (<3 ng/ml) and may therefore not be entirely generalizable. Another limitation is the lack of pre-biopsy MRI data in these studies. The ongoing ProGRESS trial is randomizing patients to either routine care or genomic-informed screening with P-CARE, with both groups strongly encouraged to undergo MRI prior to biopsy. Secondary analysis of that trial will evaluate combination and interactions between genomic risk and MRI. Another notable limitation is the early-baseline PSA analysis was limited to MVP because PCPT enrolled men aged 55 years or older, so it included no baseline PSA measurements from ages 40–49.

## Conclusions

Decisions about whom to screen, when to start screening, and whom to eventually biopsy may be informed by incorporation of germline polygenic risk with PSA. This work provides further evidence to support ongoing and planned clinical trials aimed at prospectively evaluating use of polygenic scores in clinical decision-making.

## Data availability

Full summary statistics relating to the Million Veteran Program (MVP) studies are available at dbGAP accession phs001672.

## Acknowledgments

<u>VA Million Veteran Program Core Acknowledgements</u>

MVP Program Office

- Sumitra Muralidhar, Ph.D., Program Director
US Department of Veterans Affairs, 810 Vermont Avenue NW, Washington, DC 20420
- Jennifer Moser, Ph.D., Associate Director, Scientific Programs
US Department of Veterans Affairs, 810 Vermont Avenue NW, Washington, DC 20420
- Jennifer E. Deen, B.S., Associate Director, Cohort & Public Relations
US Department of Veterans Affairs, 810 Vermont Avenue NW, Washington, DC 20420

MVP Steering Committee

- Co-Chair: J. Michael Gaziano, M.D., M.P.H.
VA Boston Healthcare System, 150 S. Huntington Avenue, Boston, MA 021
- Co-Chair: Dave Oslin, M.D.
Philadelphia VA Medical Center, 3900 Woodland Avenue, Philadelphia, PA 19104
- Sumitra Muralidhar, Ph.D., Ex-Officio
US Department of Veterans Affairs, 810 Vermont Avenue NW, Washington, DC 20420
- Drew Helmer, M.D., M.S.
Michael E. DeBakey VA Medical Center, 2002 Holcombe Boulevard, Houston, TX 77030
- Adriana Hung, M.D., M.P.H.
VA Tennessee Valley Healthcare System, 1310 24th Avenue, South Nashville, TN 37212
- Philip S. Tsao, Ph.D.
VA Palo Alto Health Care System, 3801 Miranda Avenue, Palo Alto, CA 94304
- Deepak Voora, M.D.
Durham VA Medical Center, 508 Fulton Street, Durham, NC 27705

MVP Co-Principal Investigators

- J. Michael Gaziano, M.D., M.P.H.
VA Boston Healthcare System, 150 S. Huntington Avenue, Boston, MA 02130
- Philip S. Tsao, Ph.D.
VA Palo Alto Health Care System, 3801 Miranda Avenue, Palo Alto, CA 94304

MVP Core Operations

- Jessica V. Brewer, M.P.H., Director, MVP Cohort Operations
VA Boston Healthcare System, 150 S. Huntington Avenue, Boston, MA 02130
- Kelly Cho, M.P.H, Ph.D., Director, MVP Phenomics
VA Boston Healthcare System, 150 S. Huntington Avenue, Boston, MA 02130
- Lori Churby, B.S., Director, MVP Regulatory Affairs
VA Palo Alto Health Care System, 3801 Miranda Avenue, Palo Alto, CA 94304
- Yonghui Jia, Ph.D., Director, VA Central Biorepository
VA Boston Healthcare System, 150 S. Huntington Avenue, Boston, MA 02130
- Jacob T. Kean, Ph.D., Acting Director, VA Informatics and Computing Infrastructure (VINCI)
VA Salt Lake City Health Care System, 500 Foothill Drive, Salt Lake City, UT 84148
- Saiju Pyarajan Ph.D., Director, Data and Computational Sciences
VA Boston Healthcare System, 150 S. Huntington Avenue, Boston, MA 02130
- Robert Ringer, Pharm.D., Director, VA Albuquerque Central Biorepository
New Mexico VA Health Care System, 1501 San Pedro Drive SE, Albuquerque, NM 87108
- Luis E. Selva, Ph.D., Director, MVP Biorepository Coordination
VA Boston Healthcare System, 150 S. Huntington Avenue, Boston, MA 02130
- Shahpoor (Alex) Shayan, M.S., Director, MVP PRE Informatics
VA Boston Healthcare System, 150 S. Huntington Avenue, Boston, MA 02130
- Brady Stephens, M.S., Principal Investigator, MVP Information Center Canandaigua VA Medical Center, 400 Fort Hill Avenue, Canandaigua, NY 14424
- Stacey B. Whitbourne, Ph.D., Director, MVP Cohort Development and Management
VA Boston Healthcare System, 150 S. Huntington Avenue, Boston, MA 02130

This research is based on data from the Million Veteran Program (MVP), Office of Research and Development, Veterans Health Administration, and was supported by MVP000, MVP084, and VA award I01CX002635-01 (ProGRESS). This publication does not represent the views of the Department of Veterans Affairs or the United States Government.

<u>Additional Funding</u>

This work was supported, in part, by the Prostate Cancer Foundation (23CHAL12, 24CHAL03), the U.S. Department of Defense (DOD/CDMRP PC220521), and the NIH (R01CA241410).

## Author Contributions

RK, JS, GX, and TMS conceived and designed the study, interpreted results, and drafted the manuscript. RK, JS, and GX performed data processing, statistical analyses, and generated figures and tables. RK prepared analytical workflows and provided expertise in polygenic risk modeling. TMS supervised the study and provided scientific and methodological guidance. CB, IG, RH, LK, AK, MAL, JL, BR, CT, JLV, and JW contributed to data acquisition, clinical expertise, study methodology, and/or interpretation of findings. All authors read and approved the final manuscript.

