## Supplement for "Additional value of germline genomic scores beyond prostate-specific antigen alone for long-term and near-term prostate cancer risk"

**Supplementary Data**

| **Characteristic** | **Summary** |
| --- | --- |
| Number of men | 428,512 |
| treatment | 44,092 (10.3%) |
| metastatic PCa | 6,774 (1.58%) |
| P-CARE risk groups |  |
| low | 96,068 (22.4%) |
| intermediate | 157,522 (36.8%) |
| high | 174,922 (40.8%) |
| Genetic Ancestry |  |
| European | 300,613 (70.2%) |
| African | 84,089 (19.6%) |
| Other | 43,810 (10.2%) |
| Median age [years] at: |  |
| baseline PSA | 56.4 [50.7-62.1] |
| treatment | 67.8 [62.6-72.8] |
| metastatic PCa | 72.4 [67.1-77.5] |
| last follow-up | 72.6 [65.8-77.2] |
| baseline PSA [ng/ml] | 0.9 [0.5-1.5] |

*Supplementary Table 1. Cohort characteristics for long-term risk analysis in MVP.*

| **Characteristic** | **Summary** |
| --- | --- |
| Number of biopsies | 57,706 |
| positive result | 14,896 (25.8%) |
| Number of men | 46,652 |
| P-CARE risk groups |  |
| low | 5,546 (11.9%) |
| intermediate | 13,906 (29.8%) |
| high | 27,200 (58.3%) |
| Genetic Ancestry |  |
| European | 29,993 (64.3%) |
| African | 12,440 (26.7%) |
| Other | 4,219 (9.04%) |
| Age at biopsy [years] | 65.2 [60.5-69.6] |
| PSA pre-biopsy | 5.7 [4.4-7.8] |

*Supplementary Table 2. Cohort characteristics for the near-term risk analysis in MVP.*

| **Characteristic** | **Summary** |
| --- | --- |
| Number of men | 6,054 |
| PCa diagnosis | 368 (6.08%) |
| P-CARE risk groups |  |
| low | 1,810 (29.9%) |
| intermediate | 2,760 (45.6%) |
| high | 1,484 (24.5%) |
| treatment group |  |
| placebo | 2,977 (49.2%) |
| Finasteride | 3,077 (50.8%) |
| Baseline PSA (ng/ml) | 1.1 [0.6-1.6] |
| Median age [years] at |  |
| baseline | 63 [59-67] |
| PCa diagnosis | 71 [66.5-75.2] |
| last follow-up | 83 [79.7-86.9] |

*Supplementary Table 3. Cohort characteristics for long-term risk analysis in PCPT.*

| **Characteristic** | **Summary** |
| --- | --- |
| Number of biopsies | 4,308 |
| positive result | 70 (1.62%) |
| Number of men | 4,269 |
| P-CARE risk groups |  |
| low | 1,303 (30.2%) |
| intermediate | 1,955 (45.4%) |
| high | 1,050 (24.4%) |
| treatment group |  |
| placebo | 2,115 (49.1%) |
| Finasteride | 2,193 (50.9%) |
| PSA pre-biopsy (ng/ml) | 0.9 [0.4-1.8] |
| Age at biopsy [years] | 68.9 [65.0-73.1] |

*Supplementary Table 4. Cohort characteristics for the near-term risk analysis in PCPT.*

| **Predictors** | | **Odds ratio for positive**  **biopsy result (csPCa)** |
| --- | --- | --- |
| P-CARE | mid | 1.64  [1.51-1.79] |
|  | high | 2.93  [2.69-3.18] |
| PSA pre-biopsy | | 1.97  [1.92-2.04] |
| Age at biopsy | | 1.26  [1.23-1.28] |
| Genetic Ancestry | African | 1.52  [1.44-1.60] |
|  | Other | 0.94  [0.86-1.01] |
| PRS_PSA_ | | 0.71  [0.69-0.73] |

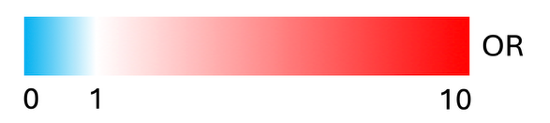

*Supplementary Table 5. Odds ratio for the near-term risk analysis in MVP limited to the first biopsy for each man. 95% confidence intervals are tabulated in square brackets. Values for PSA pre-biopsy are for every doubling in value; age at biopsy for every increase in 5 years; PRS_PSA_ for every increase in 1 standard deviation. Gray boxes denote no statistical significance.*

| **Predictors** | | **HR for csPCa**  **diagnosis** |
| --- | --- | --- |
| P-CARE | mid | 1.9  [0.92-3.92] |
|  | high | 4.62  [2.22-9.61] |
| Baseline PSA | | 2.13  [1.56-2.90] |
| Age at baseline | | 1.13  [0.92-1.38] |
| PRS_PSA_ | | 0.72  [0.56-0.92] |
| Treatment group (Finasteride) | | 0.74  [0.46-1.18] |
| Race (other) | | 1.66  [0.66-4.13] |

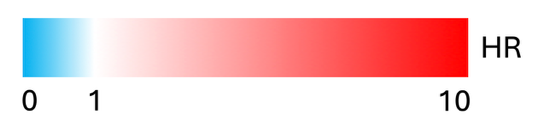

*Supplementary Table 6. Hazard ratios for csPCa in PCPT. Gray boxes denote no statistical significance.*

| **Predictors** | | **Odds ratio for positive**  **biopsy result (csPCa)** |
| --- | --- | --- |
| PHS601 | | 2.16  [2.08-2.24] |
| PSA pre-biopsy | | 1.94  [1.89-2.00] |
| age at biopsy | | 1.29  [1.27-1.31] |
| Genetic Ancestry* | African | 1.12  [1.06-1.18] |
|  | Other | 0.94  [0.87-1.01] |
| previous biopsy | | 0.23  [0.22-0.25] |
| PRS_PSA_ | | 0.7  [0.69-0.72] |

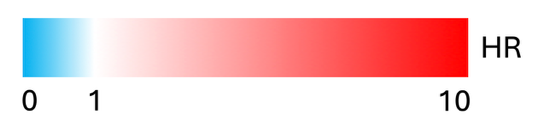

*Supplementary Table 7. Odds ratio for detection of grade group* ≥ 2 PCa on biopsy in the MVP *near-term risk analysis, PHS601 only. 95% confidence intervals are tabulated in square brackets. Values for PSA pre-biopsy are for every doubling in value; age at biopsy for every increase in 5 years; PRS_PSA_ for every increase in 1 standard deviation.* *Gray boxes denote no statistical significance. * European ancestry is reference.*

| **Characteristic** | **Long-term risk in MVP (40-49)** |
| --- | --- |
| Number of men | 92,794 |
| treatment | 6486 |
| metastatic PCa | 792 |
| P-CARE risk groups |  |
| low | 17,260 |
| intermediate | 30,727 |
| high | 44,807 |
| Genetic Ancestry |  |
| European | 49,164 |
| African | 31,352 |
| Other | 12,278 |
| Median age [years] at: |  |
| baseline PSA | 46.1 [43.5-48.3] |
| treatment | 59.9 [55.4-64.1] |
| metastatic PCa | 63.2 [58.7-66.9] |
| last follow-up | 61.7 [56.0-66.3] |
| baseline PSA [ng/ml] | 0.7 [0.5-1.1] |

*Supplementary Table 8. Cohort characteristics for long-term risk analysis in 40-49 subgroup of MVP.*

| **Baseline PSA (ng/mL)** | **P-CARE risk group** | **n** |
| --- | --- | --- |
| 0.1 to 1 | low | 516 |
|  | mid | 733 |
|  | high | 618 |
| 1 to 3 | low | 761 |
|  | mid | 1,496 |
|  | high | 1,988 |
| 3 to 10 | low | 4,968 |
|  | mid | 13,160 |
|  | high | 25,734 |
| 10 to 20 | low | 761 |
|  | mid | 2,079 |
|  | high | 4,892 |

*Supplementary Table 9. Number of MVP biopsies by pre-biopsy PSA and P-CARE risk group in the near-term analysis.*

| **Baseline PSA (ng/mL)** | **Genetic Risk Group** | **n** |
| --- | --- | --- |
| 0.1 to 1 | low | 30 |
|  | mid | 352 |
|  | high | 219 |
| 1 to 3 | low | 76 |
|  | mid | 755 |
|  | high | 401 |
| 3 to 10 | low | 673 |
|  | mid | 6,767 |
|  | high | 4,809 |
| 10 to 20 | low | 117 |
|  | mid | 1,057 |
|  | high | 886 |

*Supplementary Table 10. Number of MVP biopsies by pre-biopsy PSA and combined genetic risk group in the near-term analysis*

| **P-CARE risk group** | **PRS_PSA_** | **Genetic risk group** |
| --- | --- | --- |
| low | greater than 75^th^ percentile | low |
| intermediate | between 25^th^ and 75^th^ percentile | intermediate |
| high | less than 25^th^ percentile | high |

*Supplementary Table 11. Genetic risk groups for Figure 1. P-CARE risk groups and PRS_PSA_ values were combined into a single categorical value using the above thresholds.*

| **Baseline PSA (ng/mL)** | **P-CARE risk group** | **Estimated probability (%)** | **95% CI (%)** |
| --- | --- | --- | --- |
| 0.1–1 | Low | 0.91 | 0.80–1.01 |
|  | Intermediate | 1.65 | 1.53–1.76 |
|  | High | 3.96 | 3.74–4.19 |
| 1–3 | Low | 2.68 | 2.38–2.99 |
|  | Intermediate | 4.55 | 4.25–4.84 |
|  | High | 10.17 | 9.66–10.69 |
| 3–10 | Low | 9.57 | 8.46–10.68 |
|  | Intermediate | 15.15 | 14.08–16.22 |
|  | High | 31.19 | 29.56–32.82 |
| 10–20 | Low | 30.19 | 26.69–33.69 |
|  | Intermediate | 47.55 | 44.14–50.95 |
|  | High | 75.35 | 72.29–78.41 |

*Supplementary Table 12. Model-estimated 15-year probabilities of transition to treatment or metastasis by baseline PSA and P-CARE risk group among MVP men whose baseline PSA was measured at ages 40–49. The age-restricted analytic subgroup included 92,794 men. Probabilities and 95% confidence intervals were estimated from the multistate model.*
